# The Potential of ChatGPT as an Aiding Tool for the Neuroradiologist

**DOI:** 10.1101/2025.07.09.25331201

**Authors:** Simon Nikola, Dan Paz

**Affiliations:** Department of Radiology, Galilee Medical Center, Nahariya, Israel

**Keywords:** Artificial Intelligence, ChatGPT, Neuroradiology, CNS Tumors, Differential Diagnosis, Diagnostic Accuracy

## Abstract

**Purpose:** This study aims to explore whether ChatGPT can serve as an assistive tool for neuroradiologists in establishing a reasonable differential diagnosis in central nervous system tumors based on MRI images characteristics.

**Methods:** This retrospective study included 50 patients aged 18-90 who underwent imaging and surgery at the Western Galilee Medical Center. ChatGPT was provided with demographic and radiological information of the patients to generate differential diagnoses. We compared ChatGPT’s performance to an experienced neuroradiologist, using pathological reports as the gold standard. Quantitative data were described using means and standard deviations, median and range. Qualitative data were described using frequencies and percentages. The level of agreement between examiners (neuroradiologist versus ChatGPT) was assessed using Fleiss’ kappa coefficient. A significance value below 5% was considered statistically significant. Statistical analysis was performed using IBM SPSS Statistics, version 27.

**Results:** The results showed that while ChatGPT demonstrated good performance, particularly in identifying common tumors such as glioblastoma and meningioma, its overall accuracy (48%) was lower than that of the neuroradiologist (70%). The AI tool showed moderate agreement with the neuroradiologist (kappa = 0.445) and with pathology results (kappa = 0.419). ChatGPT’s performance varied across tumor types, performing better with common tumors but struggling with rarer ones.

**Conclusion:** This study suggests that ChatGPT has the potential to serve as an assistive tool in neuroradiology for establishing a reasonable differential diagnosis in central nervous system tumors based on MRI images characteristics. However, its limitations and potential risks must be considered, and it should therefore be used with caution.

## Introduction

Natural language processing (NLP) - the branch of artificial intelligence (AI) concerned with the interaction between computers and human language has advanced markedly in recent years with the introduction of sophisticated deep learning models.[1] Improved performance in NLP tasks such as text and speech processing have fueled impressive demonstrations of these models’ capabilities. This has captured the public imagination by seemingly imbuing computers with the ability to write “Complex, syntactically coherent thoughts.”[2] Perhaps no demonstration has been more impactful to date than with the introduction of the publicly available online chatbot ChatGPT in November 2022 by AI Lab Open Ai based on an NLP model known as a Generative Pretrained Transformer (GPT). With a simple “text in, text out” interface,[3].

ChatGPT is a very accessible language model informed by a data set that includes at least 57 billion words and 175 billion parameters from the “internet, books and other sources.”[4] The breadth of this data means that ChatGPT can handle prompts across a wide range of domains, including medicine. It can be asked to perform language-based tasks ranging from writing articles[5] to answering clinical questions,[6] although there have been complaints that it may produce seemingly credible but incorrect responses, such as that it is inventing terms that it needs to be familiar with.[7]

The Journal of the American Medical Association recently implemented guidelines for their publications regarding the use of chatbots.[8] ChatGPT potential use as a clinical tool is easy to see because it has demonstrated an ability to answer clinical questions accurately in clear and plain English, which can be understood by both healthcare providers and patients. While observing its capability, it is important to characterize its limitations. ChatGPT is only as good as its derived training data. These data are potentially biased, unreliable, and are not necessarily up to date. According to Open AI, the model has “limited knowledge of world and events after 2021.”[3,7]

The Radiology-Pathology Concordance (RPC) is an established tool for evaluation, training, and learning in medical imaging. It involves collecting data from radiology and pathology reports, comparing them, and generating a quality metric.[8] This study aims to explore whether AI tools based on NLP, specifically ChatGPT, can assist neuroradiologists in their daily work by helping to establish reasonable differential diagnoses for central nervous system tumors.

## Research Goals

The goal of this study is to assess whether AI tools based on natural language processing, particularly ChatGPT, can aid neuroradiologists in their daily work. Specifically, we aim to evaluate ChatGPT’s ability to generate reasonable differential diagnoses for central nervous system tumors and compare its performance to that of an experienced neuroradiologist.

## Materials and Methods

This retrospective study included 50 patients aged 18-90 who underwent surgery at the Galilee Medical Center. Inclusion criteria were: 1.surgery performed at the Galilee Medical Center, 2. MRI examination conducted at the center, 3. complete MRI examination including contrast injection, 4. final pathology report entered into the Chameleon system.

ChatGPT was provided with general patient data (age and gender) and relevant radiological information similar to a radiologist’s report, including 10 specific parameters.

The 10 specific parameters included were: 1. intra/extra axial location, 2. specific brain location, 3. tumor size, 4. whether the tumor is cortically based or not, 5. T1 signal characteristics, 6. T2 signal characteristics, 7. T1 post-contrast enhancement pattern, 8. presence of diffusion restriction, 9. susceptibility-weighted imaging (SWI) findings, and 10. presence of cystic components.

The AI was then asked to provide a reasonable differential diagnosis. These results were compared to the final pathology report (gold standard) and an experienced neuroradiologist’s diagnosis. Radiology and Pathology Concordance (RPC) was performed, and data were summarized in an Excel spreadsheet.

Quantitative data were described using means, standard deviations, medians, and ranges. Qualitative data were presented as frequencies and percentages. The level of agreement between examiners was assessed using Fleiss’ kappa coefficient. A p-value less than 5% was considered statistically significant. All statistical analyses were performed using IBM SPSS Statistics, version 27.

## Results

The study included 50 patients with a mean age of 59.24 years (SD = 17.19, range 21-87) (Table 1). The gender distribution was 56% female (n=28) and 44% male (n=22) (Table 2).

**TABLE 1:** Age of the patients.

| Statistics |  |  |
| --- | --- | --- |
| AGE (in years) |  |  |
| N | Valid | 50 |
|  | Missing | 0 |
| Mean |  | 59.24 |
| Median |  | 62.00 |
| Std. Deviation |  | 17.190 |
| Minimum |  | 21 |
| Maximum |  | 87 |
| Percentiles | 25 | 43.25 |
|  | 75 | 74.00 |

**TABLE 2:** Gender of the patients.

|  |  | gender |  |  | Cumulative |
| --- | --- | --- | --- | --- | --- |
|  |  | Frequency | Percent | Valid Percent | Percent |
| Valid | female | 28 | 56.0 | 56.0 | 56.0 |
|  | male | 22 | 44.0 | 44.0 | 100.0 |
|  | Total | 50 | 100.0 | 100.0 |  |

Comparing diagnoses of neuroradiologist and ChatGPT to the pathology gold standard, Neuroradiologist accuracy was 70% (35/50 correct), (Table 3).while ChatGPT accuracy was 48% (24/50 correct) (Table 4).

**TABLE 3:** Comparing diagnoses of neuroradiologist to pathology findings.

|  |  | radiologist |  |  | Cumulative |
| --- | --- | --- | --- | --- | --- |
|  |  | Frequency | Percent | Valid Percent | Percent |
| Valid | In correct | 15 | 30.0 | 30.0 | 30.0 |
|  | correct | 35 | 70.0 | 70.0 | 100.0 |
|  | Total | 50 | 100.0 | 100.0 |  |

**TABLE 4:**
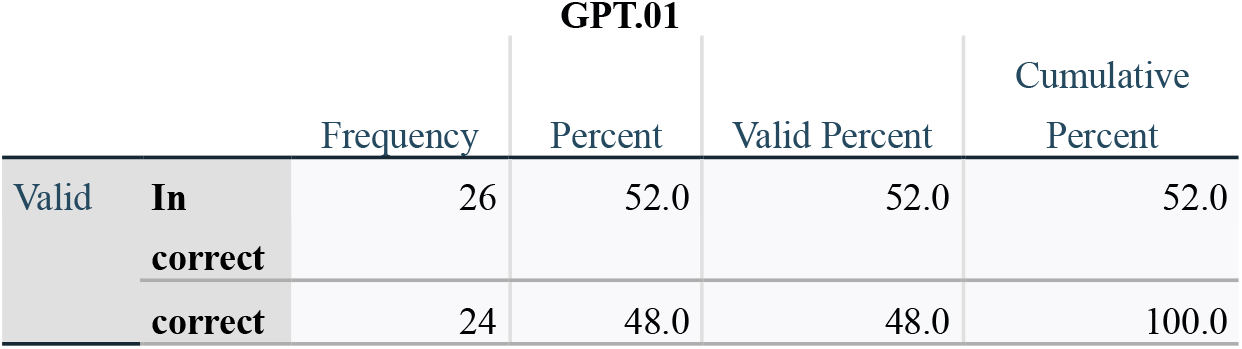
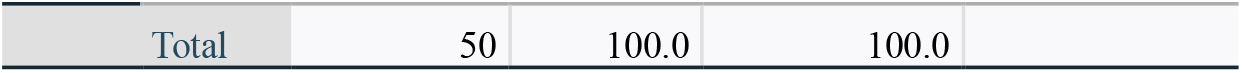
Comparing diagnoses of chatGPT to pathology findings.

The agreement between ChatGPT and the neuroradiologist was moderate (Kappa = 0.445, p<0.001) (Table 5). ChatGPT also showed moderate agreement with the pathology results (Kappa = 0.419, p<0.001) (Table 6).

**TABLE 5:** Correlation between chatGPT and neuroradiologist.

|  |  | Symmetric Measures |  |  |  |  |
| --- | --- | --- | --- | --- | --- | --- |
|  |  | Value | Asymptotic<br>Standard<br>Error<sup>a</sup> | Approximate<br>T<sup>b</sup> | Approximate<br>Significance | Exact<br>Significance |
| Measure of Agreement | Kappa | .445 | .076 | 9.040 | .000 | .000 |
| N of Valid Cases |  | 50 |  |  |  |  |

a. Not assuming the null hypothesis.
b. Using the asymptotic standard error assuming the null hypothesis.

**TABLE 6:** Correlation between GPT and pathology findings (gold standard)

|  |  | Symmetric Measures |  |  |  |  |
| --- | --- | --- | --- | --- | --- | --- |
|  |  | Value | Asymptotic<br>Standard<br>Error<sup>a</sup> | Approximate<br>T<sup>b</sup> | Approximate<br>Significance | Exact<br>Significance |
| Measure of Agreement | Kappa | .419 | .076 | 8.411 | .000 | .000 |
| N of Valid Cases |  | 50 |  |  |  |  |

a. Not assuming the null hypothesis.
b. Using the asymptotic standard error assuming the null hypothesis.

Our study included a diverse array of central nervous system neoplasms, with 25 distinct pathological entities identified. Among this spectrum of tumors, glioblastoma and meningioma emerged as the most frequent diagnosis across all three assessment modalities: neuroradiologist, ChatGPT, and histopathology.

Detailed analysis of tumor types revealed that of the cases ChatGPT thought to be meningiomas, it correctly diagnosed 6 out of 10, while the neuroradiologist correctly diagnosed all 7 of the cases they thought to be meningiomas. For cases thought to be glioblastoma, ChatGPT correctly diagnosed 10 out of 16, while the neuroradiologist correctly diagnosed 10 out of 13. Regarding cases thought to be metastases, ChatGPT correctly diagnosed 2 out of 5, while the neuroradiologist correctly diagnosed 6 out of 7. It’s important to note that these figures represent the accuracy for each tumor type as identified by ChatGPT or the neuroradiologist, not the total number of actual cases of each tumor type as determined by histopathology. (Tables 7 and 8).

**TABLE 7:**
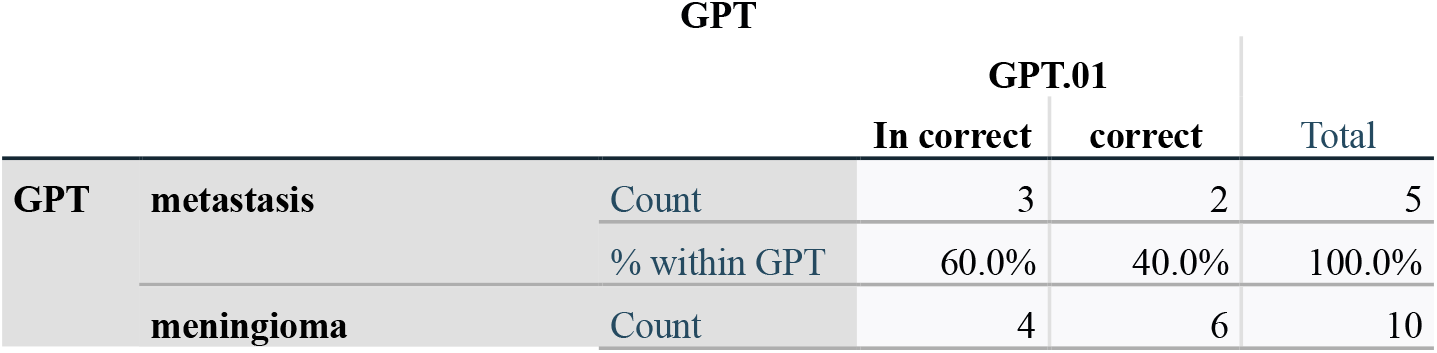
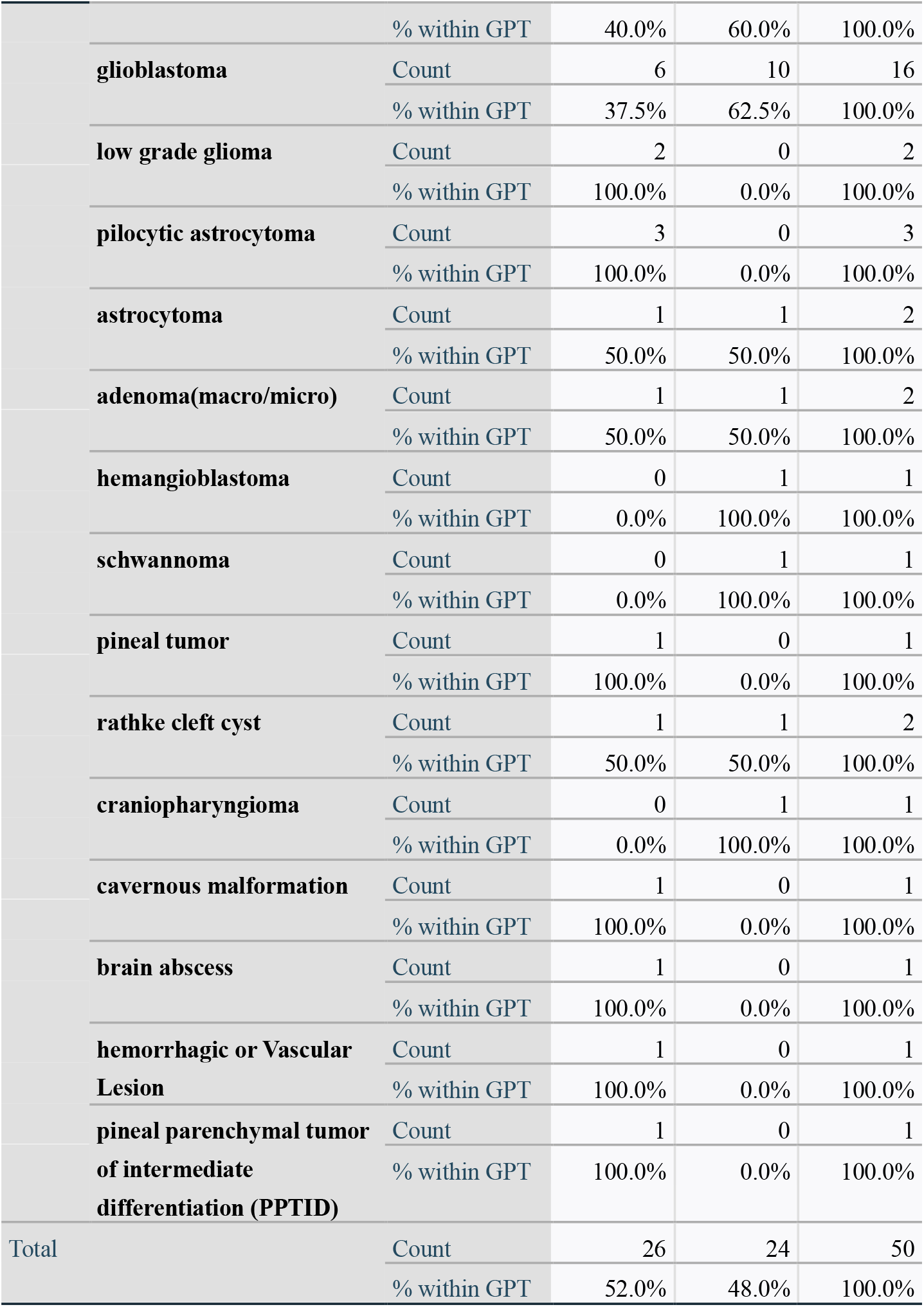
For each type of neoplasm, in how many cases did chatGPT correctly or in correctly diagnose.

**TABLE 8:** For each type of neoplasm, in how many cases did the neuroradiologist correctly or in correctly diagnose.

|  |  |  | radiologist |  | Total |
| --- | --- | --- | --- | --- | --- |
|  |  |  | In correct | correct |  |
| radiologist | **metastasis** | Count | 1 | 6 | 7 |
|  |  | % within radiologist | 6.7% | 17.1% | 14.0% |
|  | **meningioma** | Count | 0 | 7 | 7 |
|  |  | % within radiologist | 0.0% | 20.0% | 14.0% |
|  | **glioblastoma** | Count | 3 | 10 | 13 |
|  |  | % within radiologist | 20.0% | 28.6% | 26.0% |
|  | **high grade glioma** | Count | 1 | 0 | 1 |
|  |  | % within radiologist | 6.7% | 0.0% | 2.0% |
|  | **low grade glioma** | Count | 3 | 1 | 4 |
|  |  | % within radiologist | 20.0% | 2.9% | 8.0% |
|  | **astrocytoma** | Count | 0 | 1 | 1 |
|  |  | % within radiologist | 0.0% | 2.9% | 2.0% |
|  | **lymphoma** | Count | 2 | 2 | 4 |
|  |  | % within radiologist | 13.3% | 5.7% | 8.0% |
|  | **adenoma(macro/micro)** | Count | 2 | 3 | 5 |
|  |  | % within radiologist | 13.3% | 8.6% | 10.0% |
|  | **germinoma** | Count | 1 | 0 | 1 |
|  |  | % within radiologist | 6.7% | 0.0% | 2.0% |
|  | **hemangioblastoma** | Count | 0 | 1 | 1 |
|  |  | % within radiologist | 0.0% | 2.9% | 2.0% |
|  | **autoimmune encephalitis** | Count | 1 | 0 | 1 |
|  |  | % within radiologist | 6.7% | 0.0% | 2.0% |
|  | **oligodendroglioma** | Count | 0 | 1 | 1 |
|  |  | % within radiologist | 0.0% | 2.9% | 2.0% |
|  | **schwannoma** | Count | 0 | 1 | 1 |
|  |  | % within radiologist | 0.0% | 2.9% | 2.0% |
|  | **pineal tumor** | Count | 1 | 0 | 1 |
|  |  | % within radiologist | 6.7% | 0.0% | 2.0% |
|  | **craniopharyngioma** | Count | 0 | 1 | 1 |
|  |  | % within radiologist | 0.0% | 2.9% | 2.0% |
|  | **medulloblastoma** | Count | 0 | 1 | 1 |
|  |  | % within radiologist | 0.0% | 2.9% | 2.0% |
| Total |  | Count | 15 | 35 | 50 |
|  |  | % within radiologist | 100.0% | 100.0% | 100.0% |

For less common tumors, ChatGPT’s performance varied. It identified one case of each pineal tumor, hemangioblastoma, schwannoma, and craniopharyngioma. However, it also incorrectly classified 3 cases as pilocytic astrocytoma, which were not identified as such by either the radiologist or histopathology.(Table 7). The neuroradiologist showed better performance across a wider range of tumor types. For lymphomas, the radiologist classified 4 cases as lymphoma, but only 2 of these were confirmed to be lymphoma by histopathology. In the case of adenomas, 5 were classified by the radiologist, with 3 being correctly diagnosed. The radiologist also classified one case of medulloblastoma, one case of hemangioblastoma, one oligodendroglioma, one schwannoma, and one craniopharyngioma with all being correctly diagnosed by the radiologist (Table 8).

When categorizing tumors into three groups (meningioma, glioblastoma, and others), aiming to enhance the statistical power and interpretability of our analysis, allowing us to focus on the two most common tumor types while maintaining a sufficiently large sample size for the third category, ChatGPT correctly identified 60% of meningiomas, 62.5% of glioblastomas, and 33.3% of other tumors (Table 9). The neuroradiologist correctly identified 100% of meningiomas, 76.9% of glioblastomas, and 60% of other tumors (Table 10).

**TABLE 9:** For each group, in how many cases did chatGPT correctly or in correctly diagnose.

|  |  |  | GPT.01 |  | Total |
| --- | --- | --- | --- | --- | --- |
|  |  |  | In correct | correct |  |
| **GPT.01** | **meningioma** | Count | 4 | 6 | 10 |
|  |  | % within GPT.01 | 40.0% | 60.0% | 100.0% |
|  | **glioblastoma** | Count | 6 | 10 | 16 |
|  |  | % within GPT.01 | 37.5% | 62.5% | 100.0% |
|  | **other** | Count | 16 | 8 | 24 |
|  |  | % within GPT.01 | 66.7% | 33.3% | 100.0% |
| Total |  | Count | 26 | 24 | 50 |
|  |  | % within GPT.01 | 52.0% | 48.0% | 100.0% |

**TABLE 10:**
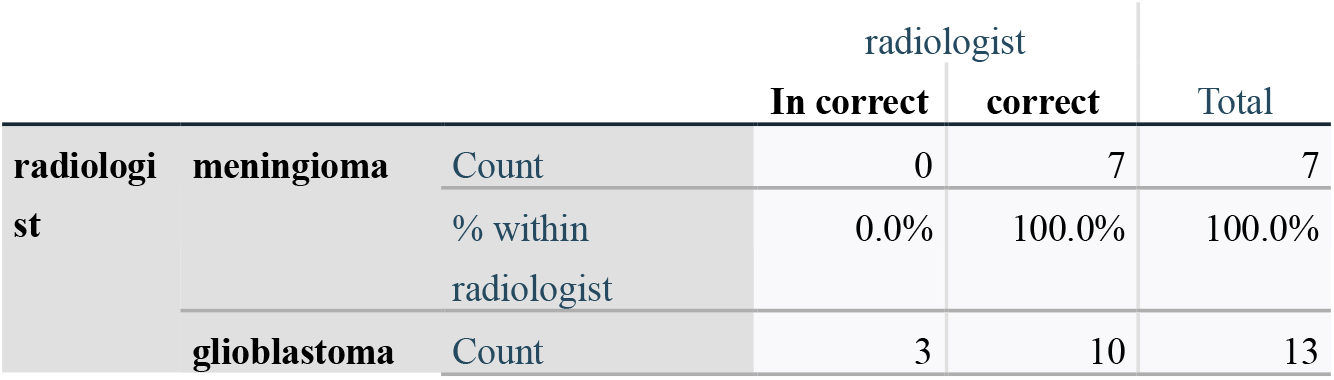
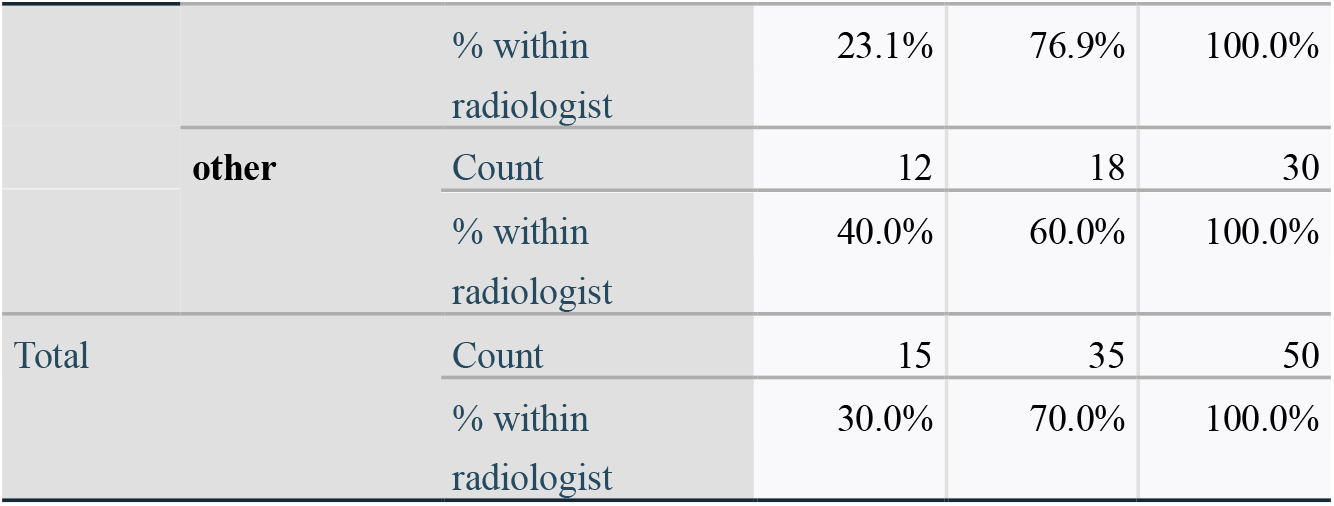
For each group, in how many cases did chatGPT correctly or in correctly diagnose.

Overall, when comparing all cases (Table 11):

**TABLE 11:** In conclusion, when comparing all cases, in how many instances were the radiologist, ChatGPT, both, or neither correct when compared to the pathology findings.

|  |  | conclusion |  |  | Cumulative |
| --- | --- | --- | --- | --- | --- |
|  |  | Frequency | Percent | Valid Percent | Percent |
| Valid | NON | 13 | 26.0 | 26.0 | 26.0 |
|  | RADIOLOGIST | 13 | 26.0 | 26.0 | 52.0 |
|  | GPT | 2 | 4.0 | 4.0 | 56.0 |
|  | BOTH | 22 | 44.0 | 44.0 | 100.0 |
|  | Total | 50 | 100.0 | 100.0 |  |

- In 44% of cases (22/50), both ChatGPT and the neuroradiologist were correct.
- In 26% of cases (13/50), only the neuroradiologist was correct.
- In 4% of cases (2/50), only ChatGPT was correct.
- In 26% of cases (13/50), neither was correct.

## Discussion

This study demonstrates that while ChatGPT shows promise in assisting neuroradiologists, its current performance falls short of human expert levels. The AI performed reasonably well in identifying common tumors like meningiomas and glioblastomas but struggled with rarer cases. This aligns with findings from other studies that have explored the use of AI in medical diagnosis [9].

The moderate agreement between ChatGPT and the neuroradiologist (Kappa = 0.445) suggests that the AI can provide valuable input, potentially serving as a “second opinion” or aiding in the initial differential diagnosis process. However, the significant gap in accuracy (48% vs 70%) underscores the continued importance of human expertise in neuroradiology, a point emphasized in recent literature on AI in healthcare [6,7].

ChatGPT’s lower performance in identifying less common tumors highlights a key limitation of current AI models. These systems are heavily dependent on the data they are trained on, and rarer conditions may be underrepresented in their training sets [4,5]. This emphasizes the need for continual updating and expansion of AI training data to improve performance across a wider range of conditions.

The study’s findings also raise important considerations about the integration of AI tools in clinical practice. While ChatGPT demonstrates potential as an assistive tool, it is clear that it should not be relied upon as a standalone diagnostic solution [7,8]. Instead, it may be most valuable when used in conjunction with human expertise, potentially helping to broaden differential diagnoses or catch rare cases that might be overlooked.

Our results support the view that large language models like ChatGPT are “double-edged swords” in medical applications [7]. While they offer significant potential benefits, their limitations and potential risks must be carefully considered. The gap between ChatGPT’s performance and that of the human expert in our study underscores the need for caution in deploying these tools in clinical settings.

The performance of ChatGPT in this study, while promising, also highlights the ongoing challenges in applying AI to complex medical tasks. As noted by Sarraju et al. [6] in their evaluation of ChatGPT for cardiovascular disease prevention recommendations, these models can provide plausible-sounding but incorrect information. This underscores the critical importance of human oversight and validation in any AI-assisted medical decision-making process.

Limitations of this study include its relatively small sample size and single-center design. Future research should expand to larger, multi-center studies to validate these findings and explore the performance of AI across diverse patient populations and clinical settings. Additionally, as AI models like ChatGPT continue to evolve rapidly [3], ongoing assessment of their capabilities and limitations in medical applications will be crucial.

## Conclusions

This study provides evidence that ChatGPT, as a representative of advanced NLP models, has potential as an assistive tool for neuroradiologists in developing differential diagnoses for central nervous system tumors. While its performance does not yet match that of experienced human experts, particularly for rare conditions, it demonstrates promise in identifying common tumors and could serve as a valuable “second opinion” tool.

The integration of AI tools like ChatGPT into clinical practice should be approached cautiously, with a clear understanding of their strengths and limitations. These tools are best viewed as supplements to, rather than replacements for, human expertise. As AI technology continues to advance, ongoing research will be crucial to assess its evolving capabilities and to develop best practices for its integration into neuroradiology and other medical fields.

Future studies should focus on larger, more diverse patient populations, explore ways to improve AI performance on rare conditions, and investigate how best to integrate these tools into clinical workflows to maximize their benefits while mitigating potential risks.

## Data Availability

All data produced in the present study are available upon reasonable request to the authors

## Notes

### Competing Interest Statement

The authors have declared no competing interest.

### Funding Statement

This study did not receive any funding

### Author Declarations

The name of the Internal Review Board (IRB)/ Approval Committee: Helsinki Committee of Galilee Medical Center Ethical approval was granted. Institutional Review Board (IRB) Approval Number: NHR-0057-23. According to the committee's decision, the following exceptions to the procedure have been approved: full exemption from informed consent.

